# Large-scale Rare Variant Burden Testing in Parkinson’s Disease Identifies Novel Associations with Genes Involved in Neuro-inflammation

**DOI:** 10.1101/2022.11.08.22280168

**Authors:** Mary B. Makarious, Julie Lake, Vanessa Pitz, Allen Ye Fu, Joseph L. Guidubaldi, Caroline Warly Solsberg, Sara Bandres-Ciga, Hampton L. Leonard, Jonggeol Jeffrey Kim, Kimberley J. Billingsley, Francis P. Grenn, Pilar Alvarez Jerez, Chelsea Alvarado, Hirotaka Iwaki, Michael Ta, Dan Vitale, Dena Hernandez, Ali Torkamani, Mina Ryten, John Hardy, UK Brain Expression Consortium (UKBEC), Sonja W. Scholz, Bryan J. Traynor, Clifton L. Dalgard, Debra J. Ehrlich, Toshiko Tanaka, Luigi Ferrucci, Thomas G. Beach, Geidy E. Serrano, Raquel Real, Huw R. Morris, Jinhui Ding, J. Raphael Gibbs, Andrew B. Singleton, Mike A. Nalls, Tushar Bhangale, Cornelis Blauwendraat

## Abstract

Parkinson’s disease (PD) has a large heritable component and genome-wide association studies to date have identified over 90 variants associated with PD, providing deeper insights into the disease biology. However, there have not been large-scale rare variant analyses for PD. To address this gap, we investigated the rare genetic component of PD at minor allele frequencies <1%, using whole genome and whole exome sequencing data from 7,184 PD cases, 6,701 proxy-cases, and 51,650 healthy controls from the Accelerating Medicines Partnership Parkinson’s disease (AMP-PD) initiative, the National Institutes of Health, the UK Biobank, and Genentech. We performed burden tests meta-analyses on protein-altering variants, prioritized based on their predicted functional impact. Our work identified several genes reaching exome-wide significance. While two of these genes, *GBA* and *LRRK2*, have been previously implicated as risk factors for PD, we identify potential novel associations for *B3GNT3, AUNIP, ADH5, TUBA1B, OR1G1, CAPN10*, and *TREML1*. Of these, *B3GNT3* and *TREML1* provide new evidence for the role of neuroinflammation in PD. To date, this is the largest analysis of rare genetic variation in PD.

## Introduction

Parkinson’s disease (PD) is a complex neurological disease likely caused by an interplay between aging, environmental factors and genetics. While the role of common genetic variation in PD has been extensively studied using large genome-wide association studies (GWAS), rare variants can also contribute to familial and sporadic disease. To date, 92 independent risk signals have been associated with PD including common variants in close proximity to *SNCA, TMEM175* and *MAPT* (Nalls et al. 2019; Foo et al. 2020). Most of the risk alleles found by array-based GWAS have frequencies over 5% in the population of interest, often reside in non-coding regions of the genome, and typically have moderate effect sizes. In contrast, rare damaging and pathogenic variants implicated in PD, such as coding variants in *SNCA* (Polymeropoulos et al. 1997) and *PRKN (Kitada et al. 1998)*, have traditionally been identified using family-based approaches. One aspect of major interest in disease genetics is the large number of pleomorphic genes, where multiple variants of varying allele frequency present with a wide range of effect sizes (A. Singleton and Hardy 2011). For example, in PD, GWAS identified common variants with moderate effects near *GBA, GCH1, LRRK2, SNCA* and *VPS13C (Nalls et al. 2019)*, while familial studies identified rare variants in the same genes resulting in more damaging effects (e.g., *GBA* p.N370S, *LRRK2* p.G2019S, and *SNCA* p.A53T) (Jansen et al. 2017; Gaare et al. 2020; Rudakou et al. 2021; Mencacci et al. 2014).

In contrast to common variation, there have been no large-scale efforts investigating the role of rare variation in PD on a genome-wide scale. Although rare variant associations for several PD genes (such as *ARSA* and *ATP10B*) have been reported in candidate gene studies (J. S. Lee et al. 2019; Martin et al. 2020), these genes remain controversial due to lack of replication in independent PD datasets (Makarious et al. 2019; Fan et al. 2020; Tesson et al. 2020; Real et al. 2020). One of the main challenges that comes with analyzing rare variants is that the quality and reliability of imputation procedures decreases with allele frequency. Since genome-wide genotyping methods are currently much cheaper than sequencing, most large datasets used for GWAS rely on imputed genotype data. A strength of the present study is that we focus on using whole genome (WGS) and whole exome sequencing (WES) to facilitate the analysis of rare variation. We perform the largest genome-wide analysis of rare variation in PD to date, investigating 7,184 PD cases, 6,701 proxy-cases (defined as having a parent or sibling with PD), and 51,650 neurologically healthy controls of European ancestry from several large sequencing efforts. Using this data, we execute gene-level burden testing in order to understand how moderate- to large-effect rare variants contribute to the genetic etiology of PD.

## Materials and Methods

### AMP-PD and NIH Genome Sequencing Data

Whole genome sequencing data was obtained from multiple datasets including the Parkinson’s Progression Markers Initiative (PPMI), the Parkinson’s Disease Biomarkers Program (PDBP), and the Harvard Biomarker Study (HBS), BioFIND, SURE-PD3, and STEADY-PD3 as part of the Accelerating Medicines Partnership in Parkinson’s Disease (AMP-PD) initiative. Several other datasets were sequenced in parallel at the Laboratory of Neurogenetics (LNG) and the U.S. Uniformed Services University (USHUS), including samples from the National Institutes of Health (NIH) PD clinic, the United Kingdom Brain Expression Consortium (UKBEC) (Trabzuni, United Kingdom Brain Expression Consortium (UKBEC), and Thomson 2014), the North American Brain Expression Consortium (NABEC) (Gibbs et al. 2010), and Wellderly (Erikson et al. 2016). All cohorts from AMP-PD (PPMI, PDBP, HBS, BioFIND, SURE-PD3, and STEADY-PD3) were processed using the GATK Best Practices guidelines set by the Broad Institute’s joint discovery pipeline and elaborated on elsewhere (Iwaki et al. 2021). All other cohorts were joint called separate from AMP-PD but in a similar manner, also from the processed WGS data following the GATK Best Practices using the Broad Institute’s workflow for joint discovery and Variant Quality Score Recalibration (VQSR)(Poplin et al. 2017). Data processing and quality control (QC) procedures have been described previously (Bandres-Ciga et al. 2020; Iwaki et al. 2021). Additional quality control was performed to exclude closely related individuals (PI_HAT >0.125) by selecting one sample at random using PLINK (v1.9; (Purcell et al. 2007)). All individuals were of European ancestry as confirmed by principal component analysis using HapMap3 European ancestry populations. Individuals recruited as part of a biased and/or genetic dataset, such as *LRRK2* and *GBA* rare variant carriers within a specific effort of PPMI, were excluded from this analysis. Including all variants within the gene boundaries, a minimum allele count (MAC) threshold of 1 was applied. Exonic regions were subset from the whole genome sequencing data using the exome calling regions from gnomAD lifted over to hg38 (Karczewski et al. 2020).

### UK Biobank

Exome sequencing data from a total of 200,643 individuals (OQFE dataset, field codes: 23151 and 23155) were downloaded from the UK Biobank in December of 2020 (Bycroft et al. 2018). Standard quality control was performed to exclude non-European outliers. Closely related individuals (PI_HAT >0.125) were excluded by selecting one sample at random using PLINK (v1.9; (Purcell et al. 2007)). Standard exome sequencing data filtering was applied using suggested parameters as described in previous UK biobank exome sequencing studies (Backman et al. 2021).

UK Biobank phenotype data were obtained from ICD10 codes (field code: 41270), PD (field code: 131023), illnesses of father and mother (field codes: 20107 and 20110), parkinsonism (field code: 42031) or dementia (field code: 42018), genetic ethnic grouping (field code: 22006), year of birth (field code: 34) and age of recruitment (field code: 21022). Cases were defined as any individual identified as having PD using the above field code. Proxy-cases were defined as having a parent or sibling with PD as previously reported (Nalls et al. 2019). Controls were filtered to exclude any individuals with an age of recruitment < 59 years, any reported nervous system disorders (Category 2406), a parent with PD or dementia (field codes: 20107 and 20110) and any reported neurological disorder (field codes: Dementia/42018, Vascular dementia/42022, FTD/42024, ALS/42028, Parkinsonism/42030, PD/42032, PSP/42034, MSA/42036).

### Genentech

Whole genome sequencing data from Genentech included a total of 2,710 PD cases and 8,994 individuals used as controls. PD cases included 2,318 individuals from 23andMe, a subset of those included in the analysis by Chang and colleagues (Chang et al. 2017) who were contacted and provided consent for this analysis. An additional 392 PD cases were obtained from the Roche clinical trial TASMAR. Individuals included as controls were obtained from various Genentech clinical trials/studies and included cases for four diseases that do not share notable heritability with PD: age-related macular degeneration (AMD, n=1,735), asthma (n=3,398), idiopathic pulmonary fibrosis (IPF, n=1,532), and rheumatoid arthritis (RA, n=2,329). Illumina HiSeq based 30× genome sequencing was performed on all samples using 150bp paired-end reads. The reads were then mapped to the GRCh38 reference genome with BWA (Li and Durbin 2009), followed by application of GATK (Li and Durbin 2009; McKenna et al. 2010) for base quality score recalibration, indel realignment, and duplicate removal. This was followed by SNP and INDEL discovery and genotyping across all samples simultaneously using variant quality score recalibration according to GATK Best Practices recommendations (DePristo et al. 2011; Van der Auwera et al. 2013; Van der Auwera and O’Connor 2020). The 11,704 samples included in these analyses passed the following QC steps: genotype missing rate < 0.1, no sample pair had kinship coefficient (k0 i.e. probability of zero alleles shared identical-by-descent; or the value Z0 reported by PLINK’s –genome module) < 0.4; and no sample was an outlier in five iterations of outlier removal using PCA (Price et al. 2006).

### Variant Annotation

Variants were annotated using the SnpEff and SnpSift annotation softwares (v4.3t; (Cingolani et al. 2012)) as well as the Ensembl Variant Effect Predictor (VEP; v104; (McLaren et al. 2016)) package. Both the Combined Annotation Dependent Depletion (CADD; v1.4; (Rentzsch et al. 2019)) and the Loss-of-Function (LoF) Transcript Effect Estimator (LOFTEE; v1.02; (Karczewski et al. 2020)) VEP plugins were used. SnpEff is a toolbox based on 38,000 genomes that is designed to annotate genetic variants and predict their downstream functional consequences. SnpSift leverages multiple databases to filter SnpEff outputs and prioritize variants, and can predict amino acid changes as having “moderate” or “high” impact. The CADD plugin for VEP is a tool used to score the deleteriousness of single nucleotide variations, insertions, and deletions. A CADD PHRED score is a scaled measure of deleteriousness, with a score of 20 indicating that the variant is among the top 1% of deleterious variants in the genome (Rentzsch et al. 2019). The LOFTEE plugin for VEP is uniquely designed to assess stop-gain, frameshift, and splice-site disrupting variants and classify these as LoF with either low or high confidence. The following variant classes were used for gene burden analyses: 1) missense variants as defined by SnpEff, 2) moderate or high impact variants as defined by SnpEff/SnpSift, 3) high confidence LoF variants as defined by LOFTEE, and 4) variants with either a CADD PHRED score > 20 or high confidence LoF variants as defined by LOFTEE.

### Gene Burden Analysis and Meta-Analysis

The AMP-PD and NIH datasets were merged prior to gene burden analysis, with duplicates removed. Rare variant testing for this merged dataset, the UK Biobank case-control dataset, and the UK Biobank proxy-control datasets were performed using the Sequence Kernel Association Test – Optimal (SKAT-O) and the Combined and Multivariate Collapsing (CMC) Wald algorithms (Seunggeun Lee, Wu, and Lin 2012; Seunggeun Lee et al. 2016). These algorithms were run using the RVtests package (v2.1.0; (Zeggini and Morris 2015)). The CMC Wald test collapses and combines all rare variants and then performs a Wald test, where only an alternative model is fit and the effect size is estimated (Seunggeung Lee et al. 2014). SKAT-O is an optimized sequencing kernel association test designed to combat limitations introduced by the SKAT and burden tests. SKAT-O aggregates the associations between variants and the phenotype of interest while allowing for SNP-SNP interactions, and has been proven to detect genes more reliably than a burden or SKAT test separately by adaptively selecting the best linear combination of both SKAT and burden tests to maximize test power (Seunggeun Lee et al. 2012). All analyses were stratified by the four variant classes described above and by maximum minor allele frequencies (MAF) levels of 1% and 0.1%. For Genentech data, SKAT-O and CMC-Wald tests were performed using the R package SKAT (Wu et al. 2011).

The combined AMP-PD and NIH dataset was adjusted for sex, age, and the first five principal components. The UK Biobank datasets were adjusted for sex, Townsend scores, and the first five principal components. For the UK Biobank analyses, only neurologically healthy controls 65 years and older were included in analyses, and therefore age was not included as a covariate. Meta-analyses of the resulting summary statistics per gene were performed using custom Python (v3.7) scripts, which we have made available on our GitHub (https://github.com/neurogenetics/PD-BURDEN). In summary, the two meta-analysis approaches used in this study were 1) a combined p-value approach using Fisher’s test, and 2) a weighted Z-score approach. In previous studies, Fisher’s method was reported to detect > 75% of causal effects (either deleterious or protective) that are in the same direction (Derkach, Lawless, and Sun 2013). Unless otherwise stated, all results reported in this manuscript correspond to the SKAT-O rare variant test, and all meta-analyses were performed using the combined p-values reported following Fisher’s test.

Rare variant analyses were performed on each dataset separately. Two joint meta-analyses were performed as follows: 1) a case-control meta-analysis between the combined AMP-PD and NIH dataset, the Genentech dataset, and the UK Biobank case-control dataset, and 2) a meta-analysis of the case-control and proxy-control results from the combined AMP-PD and NIH dataset, the Genentech dataset, the UK Biobank PD case-control dataset, the UK Biobank sibling proxy-cases dataset, and the UK Biobank parent proxy-cases dataset. A summary of the analysis workflow is outlined in **Figure 1**.

**Figure 1:**
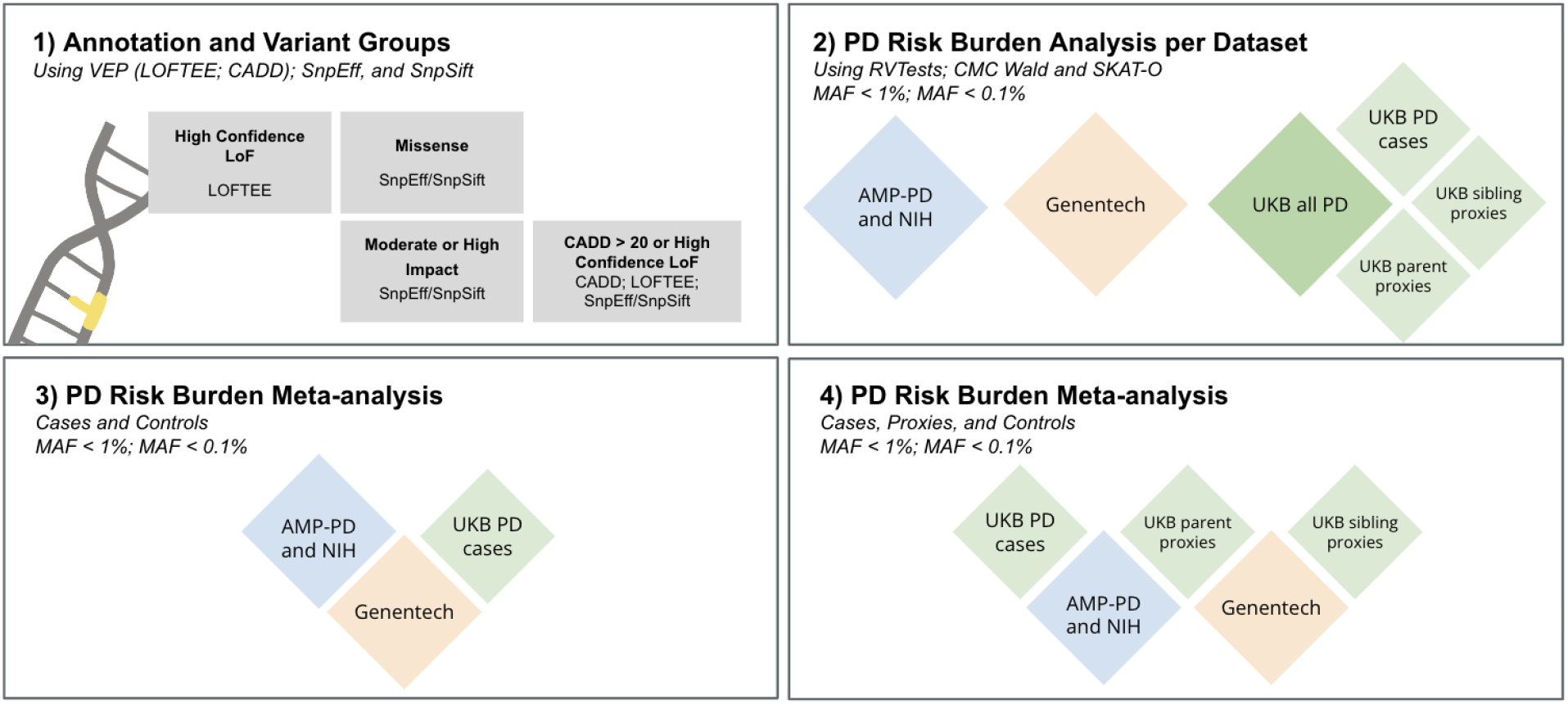
Graphical representation of the Analysis Workflow. 1) Annotation was performed using VEP and four variant groups were selected: a) missense variants as defined by SnpEff, b) moderate or high impact variants as defined by SnpEff/SnpSift, c) high confidence LoF variants as defined by LOFTEE, and d) variants with either a CADD PHRED score > 20 or high confidence LoF variants as defined by LOFTEE. 2) Burden analysis was performed on each dataset separately at rare (MAF<1%) and ultra-rare (MAF<0.1%) cut-offs. 3) Meta-analysis strategy 1 using only PD cases and controls, otherwise referred to as the “case-control” meta-analysis. 4) Meta-analysis strategy 2 using PD cases, PD proxy cases (siblings and parent), and controls, otherwise referred to as the “case-control-proxies” meta-analysis.

### Power Calculations

100 gene simulations were run using the power calculation function with default European haplotypes made available in the SKAT R package (v2.0.1; (Seunggeun Lee et al. 2012)). The total sample size was estimated at 65,535, with 7,184 PD cases, 6,701 proxy-cases down-weighted to ¼ of a PD case (corresponding to 1,675 cases), and 51,650 controls resulting in a case proportion of 13.5%. We estimated the disease prevalence of PD at 1% as previously described (Tysnes and Storstein 2017) and used an exome-wide significance threshold (α) of 1E-6. Assuming close to 20,000 protein-coding genes, resulting in a Bonferroni correction of 2.50E-6, the threshold of significance was adjusted to 1E-6 to account for two algorithms. Power calculations based on varying percentages of causality (10%, 5%, 3%, 1%, and 0.5%) and causal MAF (0.05%, 0.1%, 0.5%, 1%, 3%, and 5%) are reported in **Supplementary Table 5**. `Assuming at least 3% of the rare alleles tested are causal, this analysis has ≥ 80% power to detect associations at the tested MAF cutoffs` **(Supplementary Table 5)**.

### Data and Code Availability

All code is available on GitHub at https://github.com/neurogenetics/PD-BURDEN. AMP-PD genome sequencing data can be accessed via the AMP-PD platform https://amp-pd.org/. UK Biobank data is accessible via application at https://www.ukbiobank.ac.uk/. All gene-level summary statistics are available on the GitHub repository.

## Results

### Study overview

A total of 7,184 PD cases, 6,701 sibling/parent proxy-cases, and 51,650 controls with whole genome (AMP-PD, NIH and Genentech) or exome (UK Biobank) sequencing were included in this analysis **(Table 1)**. Rare variant gene-level burden tests were performed across all genes for four variant classes and two causal MAF cutoffs (**Figure 1**). As expected, we observed that more deleterious variant classes resulted in fewer variants tested per gene.

**Table 1:** Datasets Overview after Quality Control.

| | Sample Size | | Age<sup>^</sup> (Mean ± SD) | | Sex (Male; %) | |
| --- | --- | --- | --- | --- | --- | --- |
| Dataset | Cases | Controls | Cases | Controls | Cases sex (Male; %) | Controls sex (Male; %) |
| AMP-PD and NIH Genomes (Includes: PPMI, PDBP, HBS, BioFIND, NIH PD clinic, UKBEC, NABEC, Welllderly) | 3,369 | 4,605 | 62.1 (11.8) | 71.9 (16.2) | 63.6 | 47.6 |
| UKB case-control (WES) | 1,105 | 5,643 | 62.9 (5.24) | 64.1 (2.84) | 62.4 | 47.6 |
| UKB sibling proxy-control (WES) | 668* | 3,463 | 62.2 (5.59) | 64.1 (2.83) | 45.5 | 49.5 |
| UKB parent proxy-control (WES) | 6,033* | 28,945 | 58.1 (7.23) | 64.1 (2.82) | 42.5 | 48.7 |
| Genentech case-control (WGS) | 2,710 | 8,994 | 64.7 (10.4) | 59.2 (15.6) | 59.2 | 40.7 |
| **Total** | **7,184 cases;**<br>**6,701 proxies** | **51,650 controls** |  |  |  |  |

AMP-PD: Accelerating Medicines Partnership Parkinson's disease; NIH: National Institutes of Health; PPMI: Parkinson's Progression Markers Initiative; PDBP: Parkinson's disease Biomarkers Project; HBS: Harvard Biomarker Study; UKBEC: UK Brain Expression Consortium; NABEC: North American Brain Expression Consortium; UKB: UK Biobank
WES: Whole exome sequences; WGS: whole genome sequences
\* indicates proxy cases
<sup>^</sup>age for AMP-PD and NIH datasets reported at recruitment or baseline, ages reported for UK Biobank datasets at recruitment, ages reported for Genentech at recruitment

### Genetic burden testing in large PD case-control datasets

Initial gene burden analyses per dataset (AMP-PD and NIH Genomes, Genentech, UK Biobank cases, UK biobank sibling proxies, and UK Biobank parent proxies) resulted in several known PD genes (e.g. *GBA* and *LRRK2*) reaching significance exome-wide (P < 1E-6; **Tables 2 and 3**), confirming the validity of our approach. Lambda values per dataset showed minimal genomic inflation when adjusted for the number of cases, proxy-cases, and controls (λ_1000_; **Supplementary Table 3**). As expected, datasets with smaller sample sizes, such as the UK Biobank sibling proxy-control dataset, resulted in increased genomic deflation when analyzed separately (λ_1000_ < 0.9).

**Table 2:** Genes reaching exome-wide significance (p<1E-6) in MAF <1% in meta-analyses and individual datasets following SKAT-O.

| VARIANT CLASS (MAF <1%) | GENE | CASE ONLY META PVAL | CASE PROXIES META PVAL | AMP NIH PVAL | GNE PVAL | UKB CASE PVAL | UKB SIBLING PVAL | UKB PARENT PVAL |
| --- | --- | --- | --- | --- | --- | --- | --- | --- |
| Missense | *GBA* ** | 3.27E-14 | 1.46E-21 | 1.05E-05 | 1.32E-08 | 3.14E-04 | 0.247 | 2.15E-10 |
|  | *LRRK2* * | 7.15E-07 | 9.46E-06 | 1.96E-07 | 0.047 | 0.372 | 0.615 | 0.482 |
| Moderate or High Impact | *GBA* ** | 9.10E-15 | 1.32E-22 | 1.05E-05 | 5.70E-08 | 1.89E-05 | 0.073 | 2.15E-10 |
|  | *LRRK2* * | 7.23E-07 | 9.85E-06 | 2.09E-07 | 0.040 | 0.413 | 0.584 | 0.527 |
|  | *TUBA1B* | 0.69 | 9.02E-05 | NA | 0.647 | 0.501 | 0.352 | 9.48E-07 |
| LOF | *B3GNT3* ** | 4.40E-09 | 3.36E-09 | NA | 4.40E-09 | NA | NA | 0.032 |
|  | *CAPN10* ** | 3.60E-07 | 7.84E-07 | NA | 0.005 | 3.75E-06 | 0.053 | 0.394 |
| CADD>20<br>or LOF | *GBA* ** | 3.72E-14 | 9.12E-22 | 1.24E-05 | 6.99E-08 | 5.77E-05 | 0.130 | 2.15E-10 |
|  | *LRRK2* * | 2.49E-07 | 4.22E-06 | 2.23E-07 | 0.012 | 0.409 | 0.735 | 0.485 |
|  | *ADH5* | 4.62E-06 | 6.15E-05 | 0.512 | 0.170 | 3.13E-07 | 0.491 | 0.768 |
|  | *OR1G1* | 0.215 | 6.56E-06 | 0.848 | 0.029 | 0.620 | 6.58E-07 | 0.063 |

\* denotes genes that pass exome-wide significance ( $P < 1E-6$ ) in one meta-analysis
\*\* denotes genes that pass exome-wide significance ( $P < 1E-6$ ) in both meta-analyses

**Table 3:** Genes reaching exome-wide significance (p<1E-6) in MAF < 0.1% in meta-analyses and individual datasets following SKAT-O.

| VARIANT CLASS<br>(MAF < 0.1%) | GENE | CASE ONLY<br>META PVAL | CASE<br>PROXIES<br>META PVAL | AMP<br>NIH<br>PVAL | GNE PVAL | UKB<br>CASE<br>PVAL | UKB<br>SIBLING<br>PVAL | UKB<br>PARENT<br>PVAL |
| --- | --- | --- | --- | --- | --- | --- | --- | --- |
| Missense | *GBA* * | 1.86E-05 | 4.48E-12 | 0.022 | 2.30E-02 | 2.55E-04 | 5.41E-03 | 6.86E-08 |
| Moderate or<br>High Impact | *GBA* * | 1.71E-06 | 4.87E-16 | NA | 0.088 | 1.13E-06 | 0.001 | 5.13E-10 |
|  | *AUNIP* ** | 1.54E-08 | 2.70E-07 | NA | 0.023 | 3.04E-08 | 0.170 | 1 |
|  | *TUBA1B* | 0.690 | 9.02E-05 | NA | 0.647 | 0.501 | 0.352 | 9.48E-07 |
|  | *TREML1* * | 0.048 | 3.58E-07 | NA | 0.010 | 0.858 | 0.001 | 1.41E-05 |
| LOF | *B3GNT3* ** | 4.40E-09 | 3.36E-09 | NA | 4.40E-09 | NA | NA | 0.032 |
|  | *AUNIP* ** | 1.64E-08 | 2.04E-07 | NA | 0.024 | 3.13E-08 | 0.116 | 1 |
|  | *CAPN10* ** | 3.60E-07 | 7.84E-07 | NA | 0.005 | 3.75E-06 | 0.053 | 0.394 |
| CADD>20 or<br>LOF | *GBA* ** | 2.33E-07 | 1.20E-14 | 0.017 | 0.127 | 4.56E-07 | 8.93E-04 | 7.89E-08 |
|  | *LRRK2* | 3.46E-06 | 2.65E-06 | 0.727 | 6.15E-07 | 0.044 | 0.771 | 0.014 |
|  | *B3GNT3* | 0.334 | 0.238 | 0.264 | 0.266 | 0.460 | 1 | 0.053 |
|  | *AUNIP* * | 2.12E-07 | 1.53E-06 | 0.886 | 0.032 | 3.15E-08 | 0.125 | 1 |
|  | *OR1G1* | 0.215 | 6.56E-06 | 0.848 | 0.029 | 0.620 | 6.58E-07 | 0.063 |

\* denotes genes that pass exome-wide significance ( $P < 1E-6$ ) in one meta-analysis
\*\* denotes genes that pass exome-wide significance ( $P < 1E-6$ ) in both meta-analyses

Rare variant burden analysis of both *GBA* and *LRRK2* reached significance exome-wide in the initial analysis of missense, moderate/high impact, and LoF or highly deleterious (CADD PHRED > 20) variants. *GBA* was significant for these variant categories in both the Genentech (P=1.32E-08; P=5.70E-08; P=6.99E-08, respectively) and UK Biobank parent proxies (P=2.15E-10; P=2.15E-10; 2.15E-10, respectively) datasets. *LRRK2* was significant for these categories in the combined AMP-PD and NIH dataset (P=1.96E-07; P=2.09E-07; P=2.23E-07, respectively). LoF variants in *B3GNT3* were significant exome-wide in the Genentech dataset (P=4.40E-09) and replicated at nominal significance in the UK Biobank parent proxies dataset (P=0.032). Moderate and high impact variants in *TUBA1B* were significant in the UK Biobank parent proxies dataset (P=9.48E-07). LoF or highly deleterious variants in *ADH5* were significant in the UK Biobank cases-control dataset (P=3.13E-07), and LoF or highly deleterious variants in *OR1G1* were significant in the UK Biobank sibling proxies dataset (P=6.58E-07; **Table 2**).

Ultra-rare variant (MAF < 0.1%) burden analysis of missense, moderate/high impact, and LoF or highly deleterious variants in *GBA* were significant exome-wide in the UK biobank parent proxies dataset (P=6.88E-08; P=5.13E-10; P=7.89E-08, respectively). LoF or highly deleterious variants in *GBA* were also significant in the UK Biobank case-control dataset (P=4.56E-07). LoF or highly deleterious variants in *LRRK2* were significant in the Genentech dataset (P=6.15E-07). Moderate/high impact variants in *AUNIP* were significant in the UK Biobank case-control dataset (P=3.04E-08), and *TUBA1B* in the UK Biobank parent proxies dataset (P=9.48E-07). LoF variants in *B3GNT3* were significant in the Genentech dataset (P=4.40E-09), and *AUNIP* in the UK Biobank case-control dataset (P=3.13E-08). LoF or highly deleterious variants in *AUNIP* were significant in the UK Biobank case-control dataset (P=3.15E-08), and LoF or highly deleterious variants in *OR1G1* were significant in the UK biobank sibling proxies dataset (P=6.58E-07). Ultra-rare variant burden analysis identified no significant genes exome-wide in any of the four variant classes within the AMP-PD and NIH genomes (P < 1E-6; **Table 3**).

### Meta-analyses of large PD datasets

The first meta-analysis (herein called the case-control meta-analysis) excluded any UK Biobank proxy-cases. The second meta-analysis (herein called the case-control-proxies meta-analysis) included UK Biobank proxy-cases in addition to cases and controls. No significant divergence from expected lambda values (range: 0.97-1.00) were detected in any of the meta-analyses performed **(Supplementary Table 4)**. Rare variant burden analysis of missense, moderate/high impact, and LoF or highly deleterious variants in *GBA* were significant exome-wide across both meta-analyses ([case-control P=3.27E-14; P=9.10E-15; P=3.722E-14, respectively] and [case-control-proxies P=1.46E-21; P=1.32E-22; P=9.12E-22, respectively]). High confidence LoF variants in *CAPN10* (case-control P=3.60E-07; case-control-proxies P=7.84E-07) and *B3GNT3* (case-control P=4.40E-09; case-control-proxies P=3.36E-09) were also significant exome-wide (**Table 2**).

Ultra-rare variant burden analysis of missense and moderate/high impact variants in *AUNIP* were significant exome-wide across both meta-analyses ([case-control P=1.54E-08; P=1.64E-08, respectively] and [case-control-proxies P=2.70E-07; P=2.04E-07, respectively]). Moderate/high impact variants in *TREML1* were significant with the inclusion of proxy-cases. As in the rare variant burden analysis, ultra-rare LoF variants in *CAPN10* (case-control P=3.60E-07; case-control-proxies P=7.84E-07) and *B3GNT3* (case-control P=4.40E-09; case-control-proxies P=3.36E-09) were also significant. Notably, both rare (MAF < 1%) and ultra-rare (MAF < 0.1%) *GBA* variants showed significant associations with PD risk **(Tables 2 and 3)**.

*B3GNT3* was identified in the high confidence LoF variant class group with p-values of 4.40E-09 in the Genentech dataset and P=0.032 in the UK biobank parent proxies. However, no variants meeting this criteria were present in the AMP-PD and NIH genomes, so the association of rare LoF variation in *B3GNT3* could not be confirmed. The majority of novel candidate genes identified in this study (*B3GNT3, AUNIP, ADH5, TUBA1B, OR1G1, CAPN10*, and *TREML1*) only reached significance exome-wide using the SKAT-O test. (**Supplementary Table 8)**. Full results from the SKAT-O and CMC Wald burden tests performed for each variant class, MAF cutoff, and meta-analysis group can be found on our GitHub repository (https://github.com/neurogenetics/PD-BURDEN).

### Conditional LRRK2 analysis

Since *LRRK2* p.G2019S is a relatively common risk factor for PD, we explored whether the rare variant association at *LRRK2* is driven primarily by this variant. The observed association at *LRRK2* was lost (P > 0.05) after conditioning on the allelic status of *LRRK2* p.G2019S for all of the tested variant categories and MAF thresholds in the discovery datasets (excluding Genentech; **Supplementary Table 8)**. Besides *LRRK2* p.G2019S, no other substantial coding risk was detected.

### Assessment of previously reported PD causal or high risk genes and GWAS regions

We next attempted to replicate a large number of genes that showed rare variant associations with PD in previous studies (for a full list, please see **Supplementary Table 9**). Besides the previously discussed *GBA* and *LRRK2*, none of these genes met exome-wide significance (P > 1E-6) in our analysis. However, we did observe sub-significant association signals for LoF or highly deleterious variants in *ARSA* (P=8.73E-05) and *DNAJC6* (P=8.08E-04; **Supplementary Table 9)**. Since we did not detect a P-value of interest in *PRKN* (P=0.30), which has been robustly associated with predominantly early onset PD in previous studies, we investigated the enrichment of homozygous and potentially compound heterozygous *PRKN* mutations in PD. In the most stringent variant class (LoF or highly deleterious variants), we found a frequency of 0.41% in cases and 0.07% in controls in the combined AMP-PD and NIH dataset (**Supplementary Table 6**).

We also attempted to determine whether known PD loci identified by GWAS present rare variant associations, as has been shown previously near *SNCA, GBA, GCH1, VPS13C*, and *LRRK2* (Jansen et al. 2017; Gaare et al. 2020; Rudakou et al. 2021; Mencacci et al. 2014). We assessed a total of 80 PD GWAS regions, 78 of which were identified in the largest GWAS of Europeans (Nalls et al. 2019) and two of which were identified in the largest PD GWAS of East Asians (Foo et al. 2020). Only two genes, *GBA* and *LRRK2*, were significant after Bonferroni correction for 2,361 unique genes within 1 megabase of known PD loci, suggesting that coding variants do not play a large role in these GWAS regions, but rather that signals are driven by non-coding variation in these regions.

## Discussion

We report the results of rare variant gene burden tests of PD using the largest sample size to date including 7,184 PD cases, 6,701 proxy-cases, and 51,650 healthy controls. A meta-analysis of gene burden results reaffirms that rare variants in *GBA* and *LRRK2* are associated with PD risk in individuals with European ancestry. However, we also observed several novel PD-associated genes (*B3GNT3, AUNIP, ADH5, TUBA1B, OR1G1, CAPN10* and *TREML1*) that met exome-wide significance (P < 1E-6) in our analysis. Although these genes were not significant across all of the datasets tested (**Supplementary Table 7)**, this may be due to varied power in the different datasets due to sample size and/or geographical population differences between the datasets that influence the presence or absence of rare variants of interest. We observed the strongest evidence of a novel rare variant association at *B3GNT3*, where loss of function variation showed a significant meta-analysis P-value (P=4.40E-09) primarily driven by the Genentech (P=4.40E-09) and UK Biobank (parent proxies P=0.032) datasets. Variants meeting this criteria were not present in the combined AMP-PD and NIH genomes, requiring additional data to confirm association with PD risk. These variants in *B3GNT3* are rare, with three variants driving the association in both the Genentech and UK Biobank parent proxies datasets, and are therefore likely to be absent in the remaining datasets analyzed.

Previously suggested GWAS loci also harbor rare variants of interest, such as *SYT11, FGF20*, and *GCH1 (Pu et al. 2022)*. We identified no significant p-values in these genes, consistent with a similar, albeit smaller, analysis performed in the East Asian population (Pu et al. 2022). However, the vast majority of previously PD-associated genes were not nominated by our analysis, including *PINK1* and *PRKN (PARK2)*, which are the most common genetic cause of early onset PD (Pandey et al. 2019). This is somewhat expected since burden testing algorithms are most well-powered to detect dominant and high-risk variants such as those in *GBA* and *LRRK2*, and are less sensitive to recessive and ultra rare mutations. It is also important to note that PD patients who carry *PRKN, PINK1*, and *SNCA* mutations often have a slightly different PD phenotype (e.g. earlier onset, varying progression rates, rapid dementia onset) compared to the general PD population (Klein and Westenberger 2012). Since most PD cases included in this analysis showed onset of symptoms in their sixties, it is less likely that they will harbor pathogenic *PRKN* mutations than those with early onset PD (**Table 1)**. It is therefore likely that such mutation carriers are underrepresented in the datasets included in this study.

Immune involvement including adaptive T lymphocyte response in PD is well described and reviewed elsewhere (Mosley et al. 2012). *B3GNT3* encodes an enzyme involved in the synthesis of L-selectin required for lymphocyte homing, particularly for rolling of leukocytes on endothelial cells, facilitating their migration into inflammatory sites. *TUBA1B* encodes the 1B chain of alpha-tubulin, the main constituent of cytoskeleton. Growing evidence suggests the role of microtubule defects in progressive neuronal loss in PD (Calogero et al. 2019; Pellegrini et al. 2017). Alpha-tubulin has previously been shown to aggregate as a result of mutations in genes encoding proteins well known to be implicated in PD, including parkin (Ren, Zhao, and Feng 2003) and alpha-synuclein (Cartelli et al. 2016). *TREML1* is one of the TREM receptors that are increasingly being implicated in neurodegenerative disorders like Alzheimer’s disease, PD, and multiple sclerosis (Dardiotis et al. 2017; Feng et al. 2019; Piccio et al. 2008). *ADH5* encodes for one of the alcohol dehydrogenases, which have been studied in the past for association with PD risk with conflicting results (Kim et al. 2020; Buervenich et al. 2005; García-Martín et al. 2019). There is no clear, discernible connection between known PD biology and the function of the remaining three genes: *AUNIP, OR1G1*, and *CAPN10*. Further studies providing genetic support and functional data for these and related genes will be necessary to uncover their potential role in PD.

There are several limitations of this study. First, our analysis was restricted to individuals of European ancestry. It is important to expand rare variant analyses of PD to non-European populations as more whole genome and whole exome sequencing data becomes available. Although the sample size is large compared to previous rare variant analyses of PD, we lack power to detect associations in genes where ≤ 3% of the variants tested are putatively functional or causal, as some rare variant tests weigh rarer variants with increased penetrance and effect size differently or not at all (**Supplementary Table 5**).

Since our literature search for previously reported rare variant associations was comprehensive and not limited to late-onset PD, it is possible that failure to replicate these associations is due to our analysis focusing on associations in late-onset PD compared to controls. Additionally, our analysis included parent and sibling proxy-cases from the UK Biobank to increase statistical power. Although PD proxy-cases have shown to be valuable in large-scale studies investigating common variation (Nalls et al. 2019) and we have demonstrated their utility at detecting rare variant associations in known PD genes such as *GBA* (**Supplementary Table 7**), we acknowledge that caution should be used when searching for recessive forms of disease. Finally, the vast majority of PD patients included in this study are from the “general” PD population, of which typically less than ∼10% have a positive family history. Future rare variant studies will benefit from recruitment efforts that prioritize PD patients who are highly suspected to have a monogenic form of disease since these individuals are more likely to harbor highly pathogenic or causal mutations that have not previously been associated with PD. This strategy is being actively used for recruitment of PD patients by the Global Parkinson’s Genetics Program (Global Parkinson’s Genetics Program 2021).

Clinical heterogeneity within PD cases has been well documented (Campbell et al. 2020; Mu et al. 2017; Sauerbier et al. 2016). Analysis of rare variants restricted to subtypes of PD may identify genes important in PD subtypes but not PD as a whole. Our analysis was also restricted to SNVs and small indels. Future analyses will benefit from the use of long-read sequencing to assess the impact of structural variants, which have been shown to be important and causal for PD (A. B. Singleton et al. 2003; Scott, Chiang, and Hall 2021; Kitada et al. 1998).

Overall, we performed the largest PD genetic burden test to date. We identified *GBA* and *LRRK2* as two genes harboring rare variants associated with PD and nominated several other previously unidentified genes. Further replication in larger datasets that prioritize familial PD cases and individuals of non-European ancestry will provide greater insight into the nominated genes.

## Data Availability

Accelerating Medicines Partnership in Parkinsons Disease (AMP PD data) and quality control notebooks are access-controlled [https://amp-pd.org/], and require individual sign-up to access the data. United Kingdom Biobank (UKBiobank) data are access-controlled and require an application for access [https://www.ukbiobank.ac.uk/]. The remaining cohorts were obtained through collaborations with the National Institutes of Health (NIH) and Genentech. All data produced in the present work are contained in the manuscript. NABEC is available from NCBI dbGaP, study accession phs001300.v2.p1

https://amp-pd.org/

https://www.ukbiobank.ac.uk/

## Acknowledgements and Funding

We would like to thank all of the subjects who donated their time and biological samples to be part of this study. This research was supported in part by the Intramural Research Program of the National Institutes of Health (National Institute on Aging and National Institute of Neurological Disorders and Stroke; project numbers: 1ZIAAG00935, 1ZIANS003154, Z01-AG000949-02). This research has been conducted using the UK Biobank Resource under Application Number 33601. This study used the high-performance computational capabilities of the Biowulf Linux cluster at the National Institutes of Health (http://hpc.nih.gov).

Data used in the preparation of this article were obtained from the AMP-PD Knowledge Platform. For up-to-date information on the study, visit https://www.amp-pd.org. AMP-PD – a public-private partnership – is managed by the FNIH and funded by Celgene, GSK, the Michael J. Fox Foundation for Parkinson’s Research, the National Institute of Neurological Disorders and Stroke, Pfizer, and Verily. We would like to thank AMP-PD for the publicly available whole-genome sequencing data, including cohorts from the Fox Investigation for New Discovery of Biomarkers (BioFIND), the Parkinson’s Progression Markers Initiative (PPMI), and the Parkinson’s Disease Biomarkers Program (PDBP). The Parkinson’s Disease Biomarker Program (PDBP) consortium is supported by the National Institute of Neurological Disorders and Stroke (NINDS) at the National Institutes of Health. A full list of PDBP investigators can be found at https://pdbp.ninds.nih.gov/policy. Harvard Biomarker Study (HBS) is a collaboration of HBS investigators (full list of HBS investigators found at https://www.bwhparkinsoncenter.org/biobank) and funded through philanthropy and NIH and Non-NIH funding sources. The HBS Investigators have not participated in reviewing the data analysis or content of the manuscript.We also thank all of our Genentech colleagues involved in the Human Genetics Initiative involved in generating the sequence data including Natalie Bowers, Julie Hunkapiller, Jens Reeder, and Suresh Selvaraj. We are grateful to the Banner Sun Health Research Institute Brain and Body Donation Program of Sun City, Arizona, for the provision of human brain tissue and data. The Brain and Body Donation Program is supported by the National Institute of Neurological Disorders and Stroke (U24 NS072026 National Brain and Tissue Resource for Parkinson’s Disease and Related Disorders), the National Institute on Aging (P30 AG19610 Arizona Alzheimer’s Disease Core Center), the Arizona Department of Health Services (contract 211002, Arizona Alzheimer’s Research Center), the Arizona Biomedical Research Commission (contracts 4001, 0011, 05-901 and 1001 to the Arizona Parkinson’s Disease Consortium) and the Michael J. Fox Foundation for Parkinson’s Research. We thank the NIH NeuroBioBank (https://neurobiobank.nih.gov) for providing human brain tissue samples and data. Wellderly: This work is supported by Scripps Research Translational Institute, an NIH-NCATS Clinical and Translational Science Award (CTSA; 5 UL1TR002550). UKBEC: Consortium members include; Juan A. Botía, University of Murcia & UCL Great Ormond Street Institute of Child Health, Karishma D’Sa, Crick Institute, Paola Forabosco, Istituto di Ricerca Genetica e Biomedica, Italy, Sebastian Guelfi, Verge Genomics & UCL Great Ormond Street Institute of Child Health, Adaikalavan Ramasamy, Singapore Institute for Clinical Sciences, Regina H. Reynolds, UCL Great Ormond Street Institute of Child Health, Colin Smith, The University of Edinburgh, Daniah Trabzuni, UCL Queen Square Institute of Neurology, Robert Walker, The University of Edinburgh, Michael E. Weale, Genomics Plc, Oxford UK. This work was supported by the UK Dementia Research Institute which receives its funding from DRI Ltd, funded by the UK Medical Research Council, Alzheimer’s Society and Alzheimer’s Research UK.Medical Research Council (award number MR/N026004/1) and Medical Research Council (award number MR/N026004/1). LNG Path confirmed: We are grateful to the Banner Sun Health Research Institute Brain and Body Donation Program of Sun City, Arizona for the provision of human biological materials (or specific description, e.g. brain tissue, cerebrospinal fluid). The Brain and Body Donation Program has been supported by the National Institute of Neurological Disorders and Stroke (U24 NS072026 National Brain and Tissue Resource for Parkinson’s Disease and Related Disorders), the National Institute on Aging (P30 AG19610 Arizona Alzheimer’s Disease Core Center), the Arizona Department of Health Services (contract 211002, Arizona Alzheimer’s Research Center), the Arizona Biomedical Research Commission (contracts 4001, 0011, 05-901 and 1001 to the Arizona Parkinson’s Disease Consortium) and the Michael J. Fox Foundation for Parkinson’s Research. We thank the NIH NeuroBioBank for the provision of tissue samples. NABEC: We thank members of the North American Brain Expression Consortium (NABEC) for providing samples derived from brain tissue. Brain tissue for the NABEC cohort were obtained from the Baltimore Longitudinal Study on Aging at the Johns Hopkins School of Medicine, the NICHD Brain and Tissue Bank for Developmental Disorders at the University of Maryland, the Banner Sun Health Research Institute Brain and Body Donation Program, and from the University of Kentucky Alzheimer’s Disease Center Brain Bank. This research was supported, in part, by the Intramural Research Program of the National Institutes of Health (National Institute on Aging, National Institute of Neurological Disorders and Stroke; project numbers 1ZIA-NS003154, Z01-AG000949-02, Z01-ES101986, and UK ADC NIA P30 AG072946).

## Data Availability and Ethics Statement

Accelerating Medicines Partnership in Parkinson’s Disease (AMP PD data) and quality control notebooks are access-controlled [https://amp-pd.org/], and require individual sign-up to access the data. United Kingdom Biobank (UKBiobank) data are access-controlled and require an application for access [https://www.ukbiobank.ac.uk/]. The remaining cohorts were obtained through collaborations with the National Institutes of Health (NIH) and Genentech. Each contributing study abided by the ethics guidelines set out by their institutional review boards, and all participants gave informed consent for inclusion in both their initial cohorts and subsequent studies. The research using data from the NIH Parkinson’s Disease clinic cohort was approved by the NIH Intramural IRB under protocol number 01-N-0206. The research with the remaining cohorts was deemed “not human subjects research” by the NIH Office of IRB Operations and stated that no IRB approval is required. The NIH Intramural IRB has waived ethical approval for the overall study (IRB #001161). All data produced in the present work are contained in the manuscript. All authors and the public can access the statistical programming code used in this project for the analyses and results generation. MBM and CB take final responsibility for the decision to submit the paper for publication. NABEC is available from NCBI dbGaP, study accession phs001300.v2.p1

## Author Contributions

Concept: MBM, ABS, MAN, TB, CB

Data processing and/or Analyses: MBM, JL, VP, JLG, CWS, ABC, HLL, JJK, KJB, FPG, PAJ, HI, MT, DV, RR, HRM, JD, JRG, ABS, MAN, TB, CB

Sample/Data contributor: TB, AT, MR, JH, SWS, BJT, CLD, DJE, TT, LF, TGV, GED, ABS

Drafting of manuscript: MBM, JL, ABS, MAN, TB, CB, AYF

Final review: All

